# Quantitative Comparison of SARS-CoV-2 Nucleic Acid Amplification Test and Antigen Testing Algorithms: A Decision Analysis Simulation Model

**DOI:** 10.1101/2021.03.15.21253608

**Authors:** Phillip P. Salvatore, Melisa M. Shah, Laura Ford, Augustina Delaney, Christopher H. Hsu, Jacqueline E. Tate, Hannah L. Kirking

## Abstract

**Background:** Antigen tests for SARS-CoV-2 offer advantages over nucleic acid amplification tests (NAATs, such as RT-PCR), including lower cost and rapid return of results, but show reduced sensitivity. Public health organizations continue to recommend different strategies for utilizing NAATs and antigen tests in various settings. There has not yet been a quantitative comparison of the expected performance of these strategies.

**Methods:** We utilized a decision analysis approach to simulate the expected outcomes of six algorithms for implementing NAAT and antigen testing, analogous to testing strategies recommended by public health organizations. Each algorithm was simulated 50,000 times for four SARS-CoV-2 infection prevalence levels ranging from 5% to 20% in a population of 100000 persons seeking testing. Primary outcomes were number of missed cases, number of false-positive diagnoses, and total test volumes. Outcome medians and 95% uncertainty ranges (URs) were reported.

**Results:** Algorithms that use NAATs to confirm all negative antigen results minimized missed cases but required high NAAT capacity: 92,200 (95% UR: 91,200-93,200) tests (in addition to 100,000 antigen tests) at 10% prevalence. Substituting repeat antigen testing in lieu of NAAT confirmation of all initial negative antigen tests resulted in 2,280 missed cases (95% UR: 1,507-3,067) at 10% prevalence. Selective use of NAATs to confirm antigen results when discordant with symptom status (e.g., symptomatic persons with negative antigen results) resulted in the most efficient use of NAATs, with 25 NAATs (95% UR: 13-57) needed to detect one additional case at 10% prevalence compared to exclusive use of antigen tests.

**Conclusions:** No single SARS-CoV-2 testing algorithm is likely to be optimal across settings with different levels of prevalence and for all programmatic priorities; each presents a trade-off between prioritized outcomes and resource constraints. This analysis provides a framework for selecting setting-specific strategies to achieve acceptable balances and trade-offs between programmatic priorities and constraints.

**Disclaimer:** *The findings and conclusions in this report are those of the authors and do not necessarily represent the official position of the U*.*S. Centers for Disease Control and Prevention*.

## INTRODUCTION

The COVID-19 pandemic, caused by the SARS-CoV-2 virus, continues to cause significant morbidity, mortality, and economic hardship worldwide. Diagnostic testing is a cornerstone of COVID-19 response strategies in the U.S. and globally.^1,2^ Nucleic acid amplification tests (NAATs, such as real-time reverse transcription–polymerase chain reaction [RT-PCR]) and antigen tests are used to diagnose current infection with SARS-CoV-2 virus. NAATs are sensitive tests for SARS-CoV-2 infection and are often utilized as “gold-standard” assays for the diagnosis of COVID-19.^3^ However, programmatic implementation of NAATs may face challenges, such as long turnaround times, which hampers the ability of testing programs to be used to interrupt transmission.^4^ Additionally, NAATs often carry substantial costs associated with reagents, equipment, personnel training and salaries, and quality control Antigen tests offer several advantages over NAATs for SARS-CoV-2 testing programs, including lower costs, point-of-care administration, and rapid return of results. In particular, use of serial antigen testing may provide benefits over NAATs for controlling outbreaks in some settings, such as congregate living facilities. ^5^ To expand COVID-19 testing availability, the U.S. government distributed 150 million antigen tests in 2020.^6^

Despite the advantages of lower costs and faster turnaround time, antigen tests are generally less sensitive than NAATs for diagnosis of COVID-19, particularly for persons without COVID-19 symptoms.^3^ In many cases, it is recommended to confirm the results of either negative or positive antigen tests or both with the use of more sensitive NAATs.^5^ Several strategies for the use of antigen tests and NAATs have been recommended by public health organizations such as the U.S. Centers for Disease Control and Prevention (CDC),^5^ the World Health Organization (WHO),^7^ and the European Centre for Disease Control and Prevention (ECDC). ^8^ Depending on program goals, different strategies may be optimal for maximizing case detection, minimizing lost productivity, or minimizing the use of NAAT testing. To date, there has been no quantitative comparison of the expected performance and testing efficiency of these different strategies at various levels of prevalence. In this analysis, we evaluated the diagnostic performance and testing volumes of SARS-CoV-2 antigen and NAAT testing programs under six diagnostic algorithms using a simulation-based decision analysis approach. This activity was reviewed by CDC and was conducted consistent with applicable federal law and CDC policy.^§^

## METHODS

### Population and Model Structure

We evaluated outcomes of a modeled population of 100,000 persons seeking community-based SARS-CoV-2 testing (rather than facility-based serial testing) in settings of 5%, 10%, 15%, and 20% prevalence of SARS-CoV-2 infection. (Numerical results summarized in the text focus on the 10% prevalence level for conciseness.) Prevalence levels can vary substantially over time and geographically,^9^ and these levels of prevalence were selected as representative of the range of percent positivity by RT-PCR in a majority of U.S. states in March, 2021.^10^ Model input parameter estimates were derived from antigen test evaluations in the U.S. from September to December 2020 (Table 1). Because these primary data were collected within U.S. populations, this analysis represents expected outcomes in a U.S. setting.

**Table 1.**
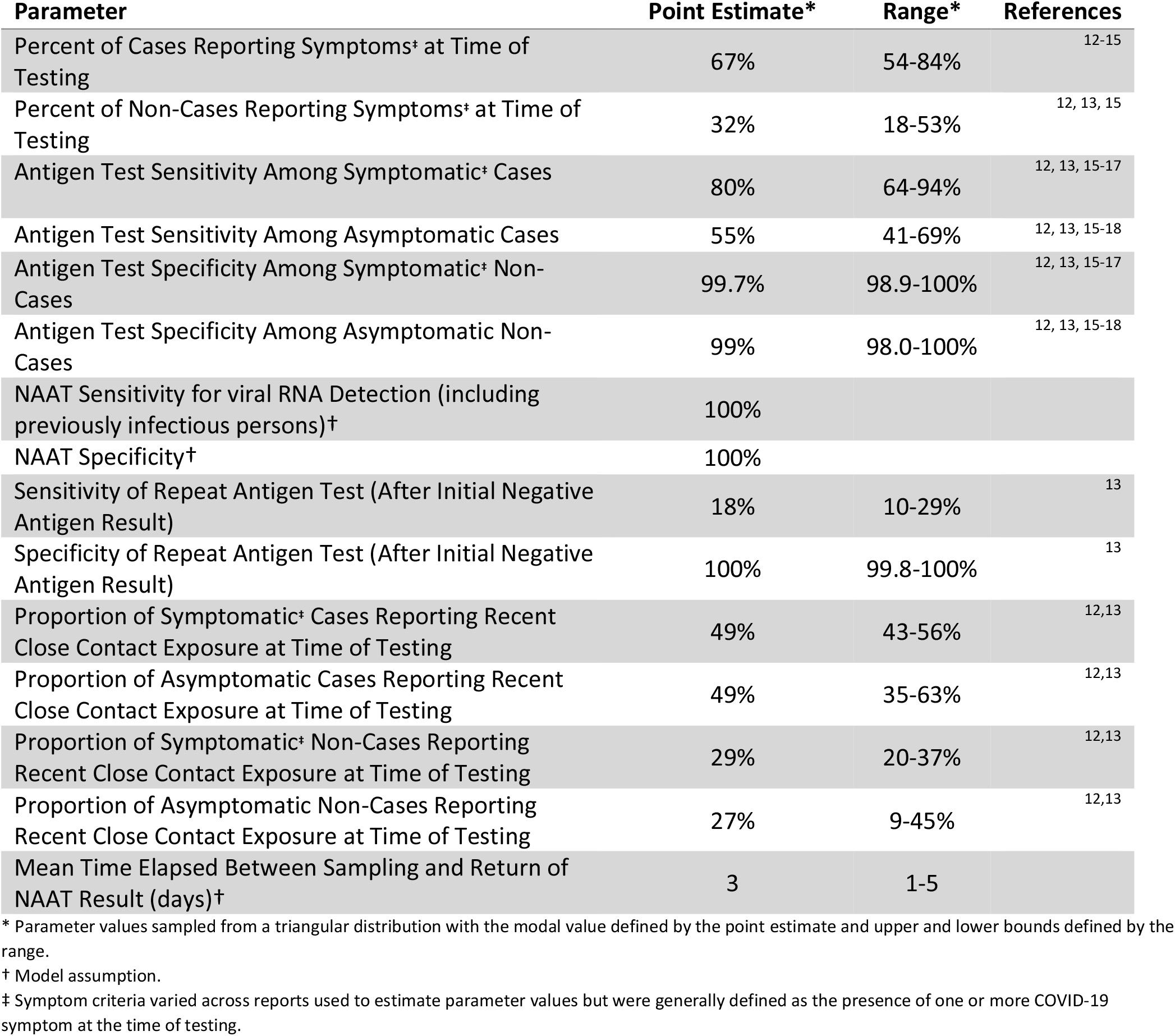
Sampling Distributions from Empiric Studies for Model Input Parameters.

| Parameter | Point Estimate* | Range* | References |
| --- | --- | --- | --- |
| Percent of Cases Reporting Symptoms<sup>‡</sup> at Time of Testing | 67% | 54-84% | 12-15 |
| Percent of Non-Cases Reporting Symptoms<sup>‡</sup> at Time of Testing | 32% | 18-53% | 12, 13, 15 |
| Antigen Test Sensitivity Among Symptomatic<sup>‡</sup> Cases | 80% | 64-94% | 12, 13, 15-17 |
| Antigen Test Sensitivity Among Asymptomatic Cases | 55% | 41-69% | 12, 13, 15-18 |
| Antigen Test Specificity Among Symptomatic<sup>‡</sup> Non-Cases | 99.7% | 98.9-100% | 12, 13, 15-17 |
| Antigen Test Specificity Among Asymptomatic Non-Cases | 99% | 98.0-100% | 12, 13, 15-18 |
| NAAT Sensitivity for viral RNA Detection (including previously infectious persons)<sup>†</sup> | 100% |  |  |
| NAAT Specificity<sup>†</sup> | 100% |  |  |
| Sensitivity of Repeat Antigen Test (After Initial Negative Antigen Result) | 18% | 10-29% | 13 |
| Specificity of Repeat Antigen Test (After Initial Negative Antigen Result) | 100% | 99.8-100% | 13 |
| Proportion of Symptomatic<sup>‡</sup> Cases Reporting Recent Close Contact Exposure at Time of Testing | 49% | 43-56% | 12,13 |
| Proportion of Asymptomatic Cases Reporting Recent Close Contact Exposure at Time of Testing | 49% | 35-63% | 12,13 |
| Proportion of Symptomatic<sup>‡</sup> Non-Cases Reporting Recent Close Contact Exposure at Time of Testing | 29% | 20-37% | 12,13 |
| Proportion of Asymptomatic Non-Cases Reporting Recent Close Contact Exposure at Time of Testing | 27% | 9-45% | 12,13 |
| Mean Time Elapsed Between Sampling and Return of NAAT Result (days)<sup>†</sup> | 3 | 1-5 |  |

\* Parameter values sampled from a triangular distribution with the modal value defined by the point estimate and upper and lower bounds defined by the range.
† Model assumption.
‡ Symptom criteria varied across reports used to estimate parameter values but were generally defined as the presence of one or more COVID-19 symptom at the time of testing.

We evaluated six diagnostic algorithms which were adapted from current recommendations for SARS-CoV-2 antigen testing in various settings. These algorithms are illustrated in Figure 1 and can be summarized as follows: *(A) NAAT Only* – each person is tested for SARS-CoV-2 infection by a NAAT; this algorithm represents an idealized scenario of unlimited NAAT capacity. *(B) Antigen (Ag) Only* – each person is tested using a single antigen test, the result of which is used as a definitive diagnosis. This algorithm represents settings with access to point-of-care antigen tests, but no access to NAAT and is analogous to interim WHO recommendations for settings with high negative predictive values.^7^ *(C) NAAT Confirmation for Symptomatic Antigen-Negative (Sx/Ag-neg) and Asymptomatic Antigen-Positive (Asx/Ag-pos) Results* – each person receives an antigen test and NAAT is used to confirm diagnoses in persons for whom antigen results do not match binary symptom status (e.g., a symptomatic person whose antigen result is negative); this algorithm represents interim U.S. CDC guidance for use of antigen tests.^5^ *(D) NAAT Confirmation of Negative Antigen Results (Ag-neg)* – each person receives an antigen test and NAAT is used to confirm negative antigen test results; this approach is analogous to ECDC options for antigen testing in high prevalence settings (≥10%).^8^ *(E) Repeat Antigen Confirmation of (Ag-neg)* – each person receives an antigen test and, for those with initial negative results, a repeat antigen test (performed within approximately 30 minutes of the initial test) is used to confirm negative diagnoses; this approach is analogous to interim WHO recommendations for settings with high prevalence.^7^ *(F) NAAT for Asymptomatic Persons (Asx) & Symptomatic Persons with Positive Antigen Results (Sx/Ag-pos)* – asymptomatic persons receive a NAAT; symptomatic persons receive an antigen test followed by a NAAT for those with positive antigen results; this approach is analogous to ECDC options for low prevalence settings and limited RT-PCR capacity.^8^

**Figure 1.**
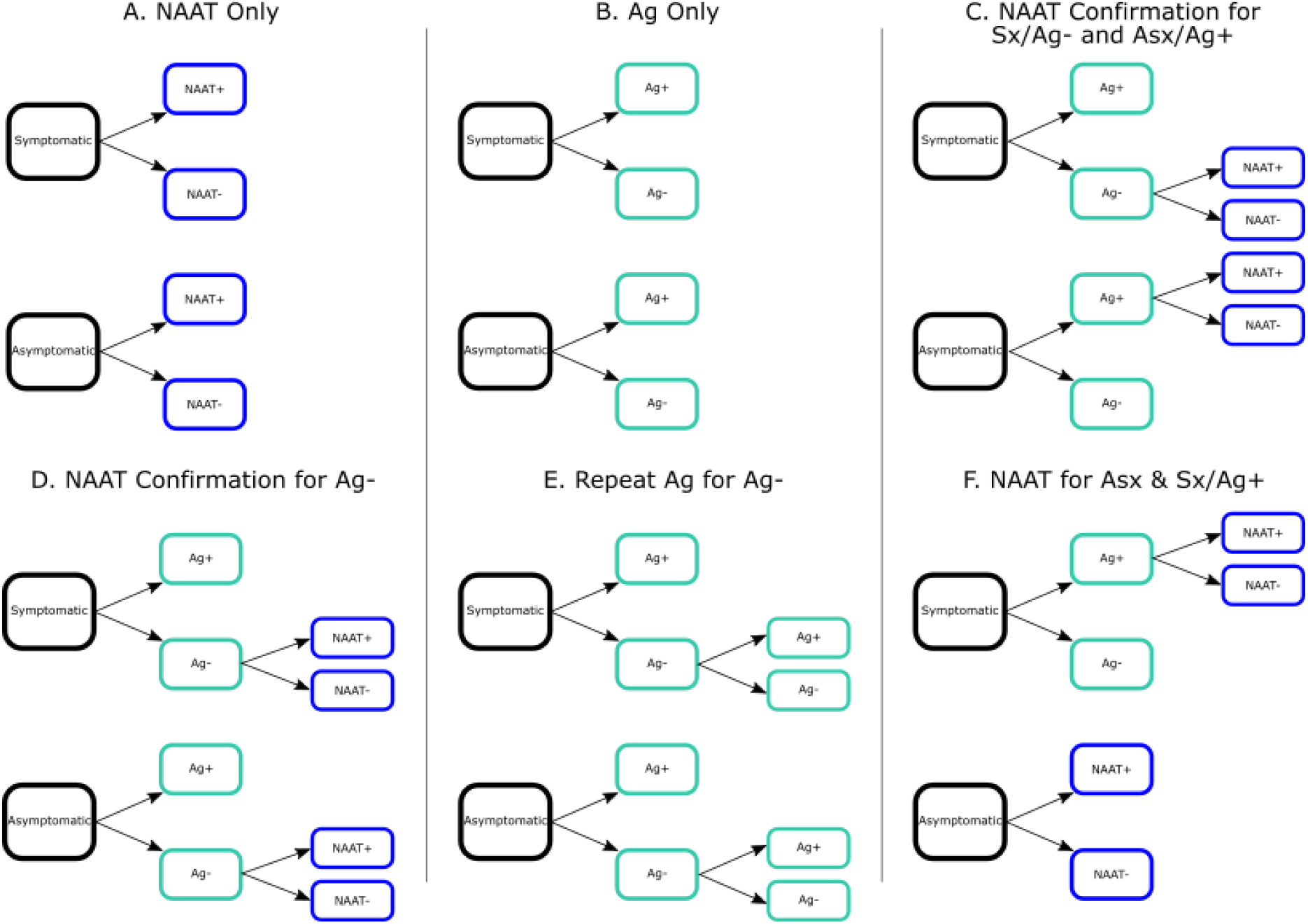
Modeled Algorithms for SARS-CoV-2 NAAT and Antigen Testing. Each panel illustrates the testing strategy utilized for one of the modeled algorithms. Algorithm abbreviations and descriptions – *(A) NAAT Only*: each person tested receives a NAAT (such as an RT-PCR test); *(B) Ag Only*: each person tested a single antigen test; *(C) NAAT Confirmation for Sx/Ag-neg and Asx/Ag-pos*: each person receives an antigen test and NAAT is used to confirm diagnoses in persons for whom antigen results do not match binary symptom status (e.g., a symptomatic person whose antigen result is negative); *(D) NAAT Confirmation of Ag-*neg: each person receives an antigen test and NAAT is used to confirm negative antigen test results; *(E) Repeat Ag Confirmation of Ag-*neg: each person receives an antigen test and, for those with initial negative results, a repeat antigen test (performed within approximately 30 minutes of the initial test) is used to confirm negative diagnoses; *(F) NAAT for Asx & Sx/Ag-pos*: – asymptomatic persons receive a NAAT, while symptomatic persons receive an antigen test followed by a NAAT for those with positive antigen results.

### Parameterization and Sampling

Parameters from empirical studies used for model simulations are summarized in Table 1. Antigen test sensitivity and specificity were assumed to be conditional on the binary symptom status of the person evaluated (symptom criteria varied across reports but was generally defined as the presence of one or more COVID-19 symptom^11^ at the time of testing) and representative of mixed populations of adults and children seeking community-based testing; the prevalence of symptoms was modeled independently for infected and uninfected populations. We made the parsimonious assumption that sensitivity and specificity of NAATs are 100% as NAATs are typically considered the “gold standard” for diagnosis of SARS-CoV-2 infection; this assumption simplifies comparison across algorithms, but may not fully represent the complex dynamics of RT-PCR positivity throughout a course of infection. Sensitivity and specificity of repeat antigen testing were assumed to be conditional upon negative initial antigen results. For the purposes of estimating lost productivity, persons evaluated were modeled to have indications for quarantine (for persons exposed to SARS-CoV-2 but testing negative)/isolation (for persons diagnosed with SARS-CoV-2 infection) based on the proportion of persons reporting recent close contact exposure at the time of testing (see below for quarantine criteria). A simplifying assumption was made for perfect sensitivity and specificity of NAATs in identifying SARS-CoV-2 infection.

Parameters were sampled from triangular distributions (defined by a modal value and upper/lower bounds, characterized in Table 1) using Latin hypercube sampling to generate 50,000 simulations of each algorithm at each prevalence level. Triangular distributions were selected as a parsimonious approach given evolving evidence from empirical studies for antigen testing characteristics. Outcomes are reported as the median and 95% uncertainty range (UR) of simulations for each scenario. All calculations and analyses were performed using R software version 4.0.2 (R Core Team, Vienna, Austria). Code for the algorithm simulations can be found on the CDC Epidemic Prediction Initiative GitHub site (https://github.com/cdcepi).

### Primary and Secondary Outcomes

Primary outcomes of interest were numbers of missed cases (persons with SARS-CoV-2 infection who receive a definitive diagnosis of “uninfected” by antigen testing with no recommendation for additional testing), false positive diagnoses (uninfected persons with a definitive diagnosis of “infected” by antigen testing with no recommendation for additional testing), and numbers of antigen tests and NAATs performed per 100,000 persons evaluated.

To quantify potential lost productivity due to longer turnaround times associated with NAATs, a secondary outcome of interest was the number of person-days of unnecessary quarantine/isolation incurred while awaiting NAAT results in each scenario. Evaluated persons were assumed to have indications for quarantine/isolation while awaiting NAAT results for the following criteria: recent close contact exposure at the time of testing; initial Ag+ results; or presence of symptoms without antigen testing results [e.g., in *(A) NAAT Only* or *(F) NAAT Confirmation for Asx & Sx/Ag-pos* algorithms]. All persons meeting these criteria were assumed to isolate/quarantine for each simulation’s sampled NAAT turnaround time in days (see Table 1).

### Incremental Outcomes and Trade-Off Analysis

To characterize the potential consequences of adopting different testing algorithms in settings of varying NAAT capacity, we calculated [compared to the *(A) NAAT Only* algorithm] the incremental number of missed cases and saved NAATs [how many fewer NAATs were needed relative to *(A) NAAT Only*] under each algorithm. These incremental measures, calculated as a quotient representing the number of NAATs saved for each additional missed case compared to the *(A) NAAT Only* algorithm, provide an indication of the number of NAATs saved under different algorithms and the consequent trade-off of additional missed cases.

A similar incremental outcome was evaluated by comparing different testing algorithms to the *(B) Ag Only* algorithm and calculating the number of additional NAATs needed and consequent trade-off of additional cases detected [compared to *(B) Ag Only* testing]. These measures are also presented as a quotient representing the number of additional NAATs needed for each additional case detected.

### Sensitivity Analyses

To identify the parameters most strongly influencing the number of missed cases under each testing algorithm, multivariable nonparametric partial rank correlation coefficients (PRCCs) were calculated to quantify the strength of correlation between individual parameter values and the number of missed cases (adjusting for all other parameter values) across the 50,000 simulations of the testing algorithm at 5% prevalence. (Monotonicity between parameter values and missed cases was verified for each parameter.) *(A) NAAT Only* and *(D) NAAT Confirmation of Ag-neg* algorithms were excluded from sensitivity analyses as they result in zero missed cases (Supplementary Figure S1). Based on these results, we performed two-way sensitivity analyses between the prevalence of symptoms among infected persons and the sensitivity of antigen tests among symptomatic cases and, separately the sensitivity of antigen tests among asymptomatic cases (Supplementary Figure S2). For two-way sensitivity analyses, the two parameters of interest are varied independently across their ranges while all other parameters are simulated at their (constant) modal values.

## RESULTS

### Primary Outcomes

Primary outcomes for each algorithm are presented in Figure 2, for settings with SARS-CoV-2 prevalence ranging from 5% to 20% among 100,000 persons evaluated. (Detailed results are available in Supplementary Table S1.) Across prevalence levels, missed cases were greatest for algorithms that did not confirm negative antigen results with NAATs, *(B) Ag Only* and *(E) Repeat Ag for Ag-neg*. At 10% prevalence, these algorithms resulted in 2,830 missed cases [*(B) Ag Only* 95% UR: 1,890-3,740] and 2,280 missed cases [*(E) Repeat Ag for Ag-neg* 95% UR: 1,507-3,067], respectively. Algorithms in which NAATs were performed prior to all definitive negative diagnoses [*(A) NAAT Only* and *(D) NAAT Confirmation of Ag-neg*], resulted in zero missed cases (due to assumed 100% sensitivity of NAATs). The remaining algorithms in which some but not all negative antigen results are confirmed by NAAT [*(C) NAAT Confirmation for Sx/Ag-pos & Asx/Ag-neg* and *(F) NAAT Confirmation for Asx & Sx/Ag-pos*], resulted in intermediate numbers of missed cases. At 10% prevalence, these algorithms result in 1,409 missed cases (95% UR: 815-2,100) and 1,389 missed cases (95% UR: 622-2,280), respectively.

**Figure 2.**
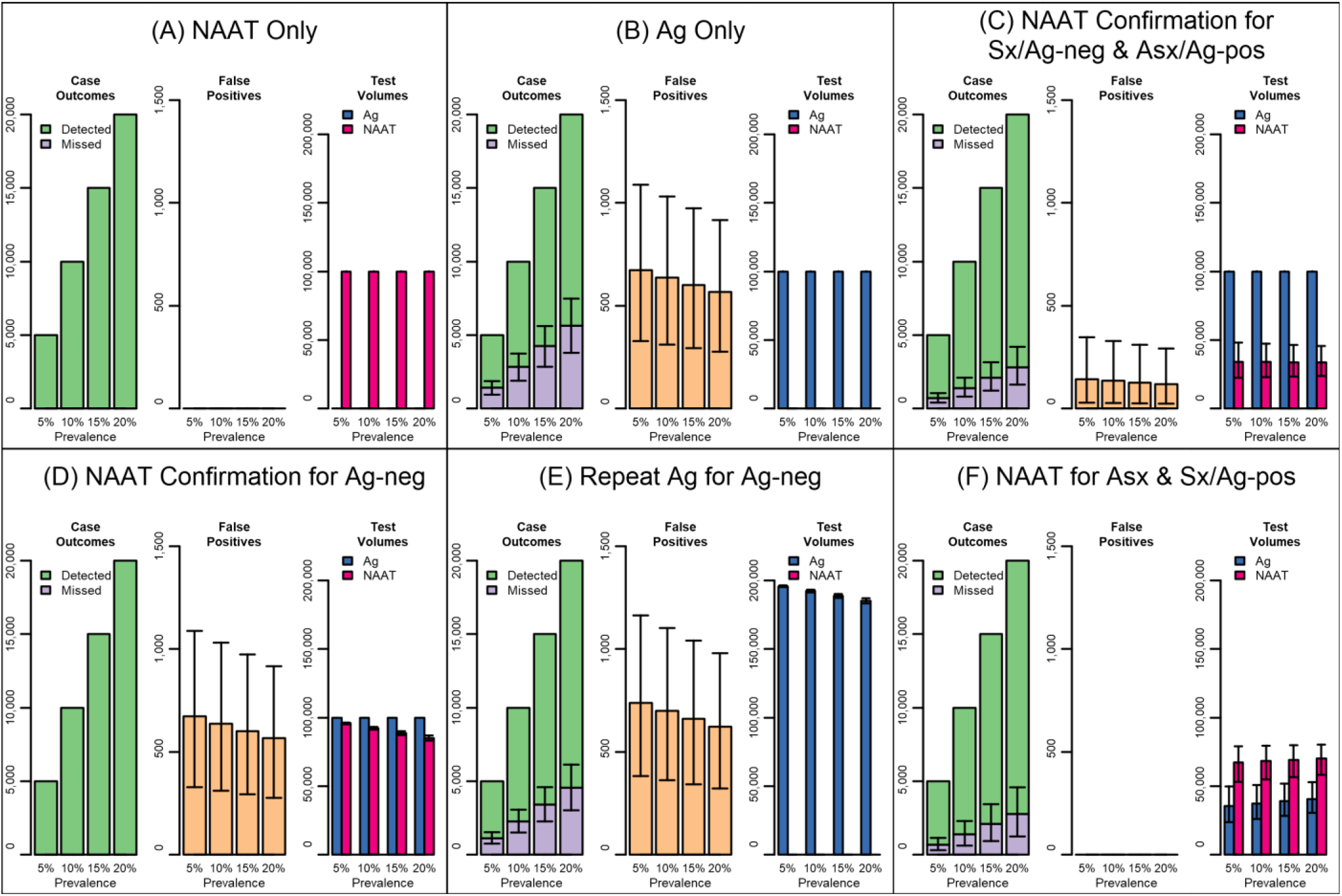
Primary Outcomes (Missed Cases, False Positives, and Test Volumes) of SARS-CoV-2 Nucleic Acid Amplification Test (NAAT) and Antigen (Ag) Testing Algorithms per 100,000 Persons Tested. Each panel presents the primary outcomes for one of the six algorithms investigated across four levels of prevalence. The left-hand graph of each panel shows the number of detected cases (in green) and missed cases (in purple). Each column of the left-hand graph sums to the total number of infected cases at each prevalence level. The middle graph of each panel shows the number of false positive diagnoses. The right-hand graph of each panel shows the number of NAATs (in magenta) and antigen tests (in blue) used. Bars represent median values and error bars represent 95% Uncertainty Ranges. Algorithm abbreviations and descriptions – *(A) NAAT Only*: each person tested receives a NAAT (such as an RT-PCR test); *(B) Ag Only*: each person tested a single antigen test; *(C) NAAT Confirmation for Sx/Ag-neg and Asx/Ag-pos*: each person receives an antigen test and NAAT is used to confirm diagnoses in persons for whom antigen results do not match binary symptom status (e.g., a symptomatic person whose antigen result is negative); *(D) NAAT Confirmation of Ag-*neg: each person receives an antigen test and NAAT is used to confirm negative antigen test results; *(E) Repeat Ag Confirmation of Ag-* neg: each person receives an antigen test and, for those with initial negative results, a repeat antigen test (performed within approximately 30 minutes of the initial test) is used to confirm negative diagnoses; *(F) NAAT for Asx & Sx/Ag-pos*: – asymptomatic persons receive a NAAT, while symptomatic persons receive an antigen test followed by a NAAT for those with positive antigen results.

False positive diagnoses were greatest in algorithms in which positive antigen results were not confirmed by NAATs— *(B) Ag Only, (D) NAAT Confirmation for Ag-neg*, and *(E) Repeat Ag for Ag-neg*. The first two of these algorithms resulted in identical numbers of false positive diagnoses (median=635 [95% UR: 311-1,031] false positive diagnoses at 10% prevalence) as both consider initial positive antigen results as definitive, while *(E) Repeat Ag for Ag-neg* resulted in higher numbers (median=699 [95% UR: 361-1,105] false positive diagnoses at 10% prevalence) due to false positive diagnoses following the repeat antigen test. Algorithms where NAATs were performed prior to all definitive positive diagnoses [*(A) NAAT Only* and *(F) NAAT Confirmation for Asx & Sx/Ag-pos*], resulted in zero false positive diagnoses (assumed 100% specificity of NAATs). The remaining algorithm [*(C) NAAT Confirmation for Sx/Ag-pos & Asx/Ag-neg*], where some but not all positive antigen results are confirmed by NAAT, resulted in low numbers of missed cases (median=134 [95% UR: 27-330] at 10% prevalence).

Total testing volume remained constant for *(A) NAAT Only* and *(B) Ag Only* algorithms, at 100,000 NAAT or antigen tests, respectively. Antigen testing also remained constant at 100,000 tests for *(C) NAAT Confirmation for Sx/Ag-neg & Asx/Ag-pos* and *(D) NAAT Confirmation for Ag-neg* algorithms. Antigen testing volume was highest for the *(E) Repeat Ag for Ag-neg* algorithm and varied depending on the number of initial negative antigen results and total volume ranged from a median of 185,100 tests (95% UR: 183,200-187,000) at 20% prevalence to a median of 195,700 tests (95% UR: 195,100-196,300) at 5% prevalence. Among algorithms using antigen testing, antigen testing volume was lowest for *(F) NAAT Confirmation for Asx & Sx/Ag-pos* and varied depending on the prevalence of symptoms among persons evaluated, ranging from a median of 35,500 tests at 5% prevalence (95% UR: 23,800-49,700) to a median of 40,700 tests at 20% prevalence (95% UR: 30,500-52,900). Among algorithms using NAATs, NAAT testing volume was lowest for *(C) NAAT Confirmation for Sx/Ag-neg & Asx/Ag-pos*: at 10% prevalence, a median of 34,100 NAATs were used (95% UR: 22,500-48,100). NAAT testing volume was higher for the *(F) NAAT Confirmation for Asx & Sx/Ag-pos* and the *(D) NAAT Confirmation for Ag-neg*: at 10% prevalence, a median of 68,300 (95% UR: 54,900-79,400) NAATs and 92,200 (95% UR: 91,200-93,200) NAATs were used, respectively.

### Secondary Outcomes: Unnecessary Quarantine

Total person-time spent in unnecessary quarantine (due to uninfected persons quarantining while waiting the return of NAAT results) is presented in Figure 3. Algorithms which did not utilize NAATs [i.e., *(B) Ag Only* and *(E) Repeat Ag for Ag-neg*] resulted in no person-days of unnecessary quarantine while awaiting NAAT results. However, these algorithms resulted in higher numbers of definitive false-positive diagnoses (as described above), which can result in up to 14 days of unnecessary isolation after the return of results per false-positive diagnosis. Among algorithms which use NAATs [*(F) NAAT Confirmation for Asx & Sx/Ag-pos*] resulted in the least person-time of unneeded quarantine, related to the low numbers of NAATs used: a median of 45,600 person-days (95% UR: 17,500-93,200) at 10% prevalence. Algorithm *(A) NAAT Only* resulted in the most person-time of unneeded quarantine, a median of 138,200 person-days (95% UR: 64,000-229,600) at 10% prevalence. However, this algorithm resulted in zero false positive diagnoses, as described above.

**Figure 3.**
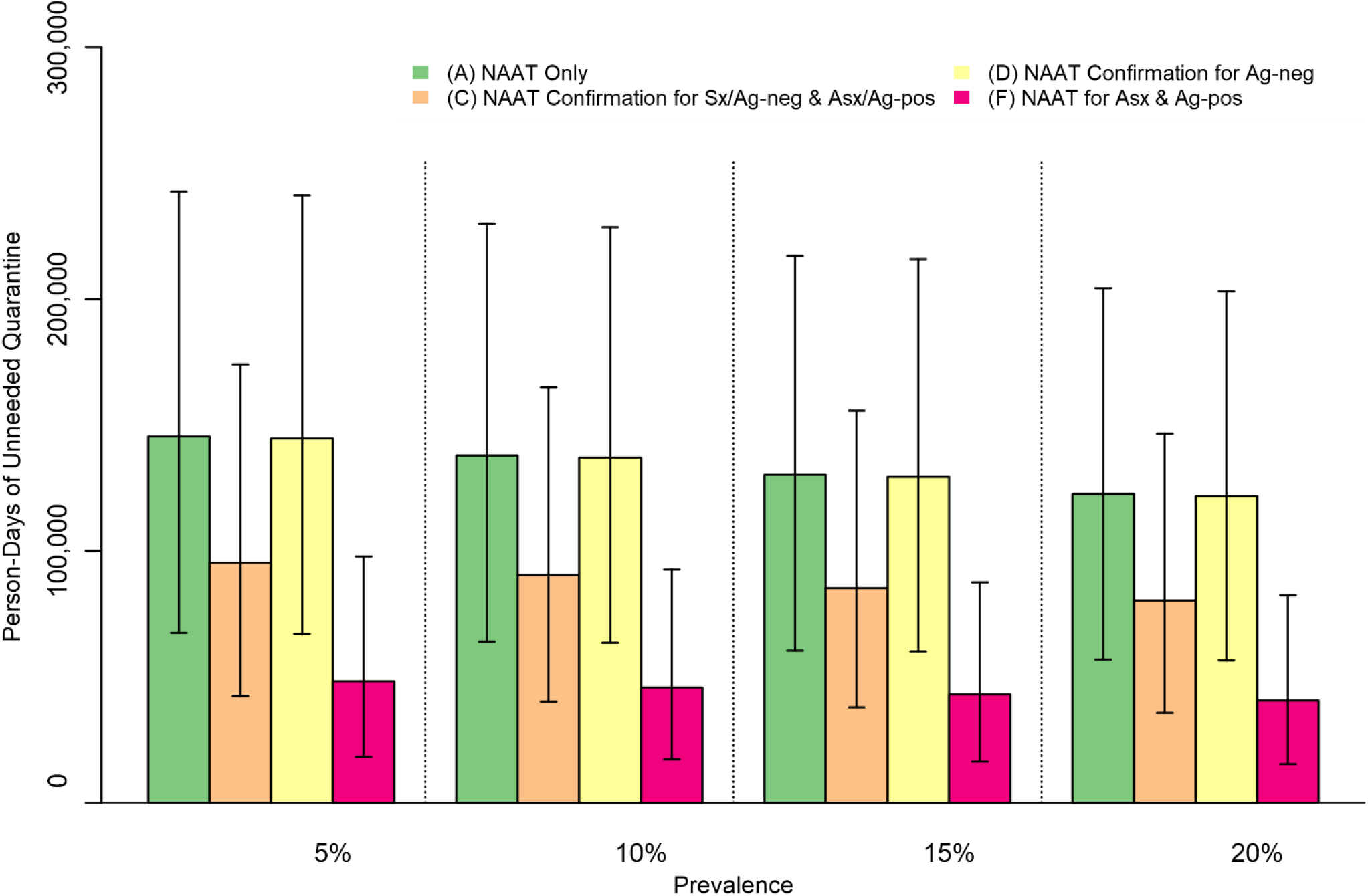
Person-Time of Unnecessary Quarantine While Waiting for NAAT Results per 100,000 Persons Tested. The total amount of person-time spent awaiting the results of NAATs by uninfected persons seeking testing are depicted at four levels of prevalence in a population of 100,000 seeking testing. Any person with a definitive antigen result and no indication for further NAAT evaluation, and asymptomatic persons with no recent close-contact exposures and no positive antigen results were excluded. Bars represent median values and error bars represent 95% Uncertainty Ranges. Algorithm abbreviations and descriptions – *(A) NAAT Only*: each person tested receives a NAAT (such as an RT-PCR test); *(B) Ag Only*: each person tested a single antigen test; *(C) NAAT Confirmation for Sx/Ag-neg and Asx/Ag-pos*: each person receives an antigen test and NAAT is used to confirm diagnoses in persons for whom antigen results do not match binary symptom status (e.g., a symptomatic person whose antigen result is negative); *NAAT Confirmation of Ag-*neg: each person receives an antigen test and NAAT is used to confirm negative antigen test results; *(E) Repeat Ag Confirmation of Ag-*neg: each person receives an antigen test and, for those with initial negative results, a repeat antigen test (performed within approximately 30 minutes of the initial test) is used to confirm negative diagnoses; *(F) NAAT for Asx & Sx/Ag-pos*: – asymptomatic persons receive a NAAT, while symptomatic persons receive an antigen test followed by a NAAT for those with positive antigen results.

### Incremental Outcomes and Trade-Offs

Incremental outcomes of simulations under algorithms compared to corresponding simulations under the *(A) NAAT Only* algorithm are depicted in Figure 4A (plotted as additional missed cases vs. NAATs saved, compared to *(A) NAAT Only*) at a level of 10% prevalence. The quotient of these measures is defined as the ratio of NAATs saved per additional missed case in Figure 4B. The *(D) NAAT Confirmation for Ag-neg* algorithm had a ratio of positive infinity, resulting from zero additional missed cases (and a small number of NAATs saved). The *NAAT Confirmation for Sx/Ag-neg & Asx/Ag-pos* algorithm had the most favorable ratio among remaining algorithms: at 10% prevalence, a median of 46 NAATs were saved per additional missed case (95% UR: 29-83) compared to *(A) NAAT Only*.

**Figure 4.**
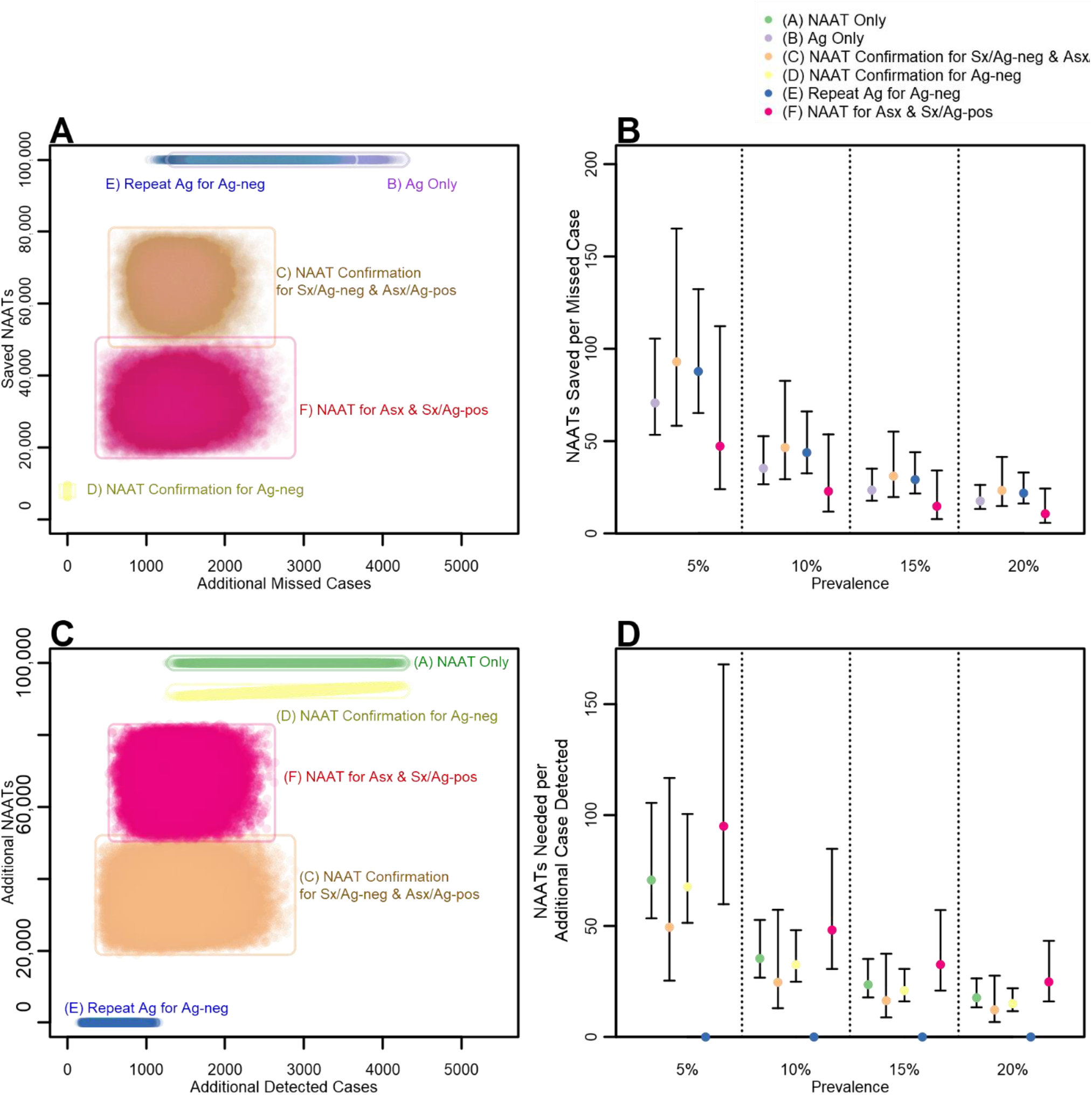
Trade-Offs in Algorithms for SARS-CoV-2 NAAT and Antigen Testing. Panel A depicts two primary outcomes (missed cases and NAAT volume) of 50,000 simulations for each of five algorithms compared to simulations of the *(A) NAAT Only* algorithm run under the same conditions at 10% prevalence in a population of 100,000 seeking testing. Panel B represents these results as a ratio of NAATs saved per missed case compared to the NAAT Only algorithm. The *(D) NAAT Confirmation for Ag-neg* algorithm results in zero missed cases, therefore this ratio equals positive infinity for all simulations and is not displayed. Panel C depicts missed cases and NAAT volume of 50,000 simulations compared to simulations of the *(B) Ag Only* algorithm run under the same conditions at 10% prevalence in a population of 100,000 seeking testing. Panel D represents these results as a ratio of NAATs needed per additional case detected compared to the Ag Only algorithm. In Panels A and C, each point represents the results of one simulation. In Panels B and D, points represent median values and error bars represent 95% Uncertainty Ranges. Algorithm abbreviations and descriptions – *(A) NAAT Only*: each person tested receives a NAAT (such as an RT-PCR test); *(B) Ag Only*: each person tested a single antigen test; *(C) NAAT Confirmation for Sx/Ag-neg and Asx/Ag-pos*: each person receives an antigen test and NAAT is used to confirm diagnoses in persons for whom antigen results do not match binary symptom status (e.g., a symptomatic person whose antigen result is negative); *(D) NAAT Confirmation of Ag-* neg: each person receives an antigen test and NAAT is used to confirm negative antigen test results; *(E) Repeat Ag Confirmation of Ag-*neg: each person receives an antigen test and, for those with initial negative results, a repeat antigen test (performed within approximately 30 minutes of the initial test) is used to confirm negative diagnoses; *(F) NAAT for Asx & Sx/Ag-pos*: – asymptomatic persons receive a NAAT, while symptomatic persons receive an antigen test followed by a NAAT for those with positive antigen results.

Incremental outcomes compared to the *(B) Ag Only* algorithm are depicted in Figure 4C (plotted as additional cases detected vs. additional NAATs needed) at 10% prevalence. These measures are presented as a ratio of additional NAATs needed per additional cases detected in Figure 4D. The *(E) Repeat Ag for Ag-neg* algorithm (which uses zero NAATs) had a ratio of zero additional NAATs needed per additional case. Among the remaining algorithms, the *(C) NAAT Confirmation for Sx/Ag-neg & Asx/Ag-pos* algorithm had the most favorable ratio: at 10% prevalence, a median of 25 NAATs were needed to detect each additional case (95% UR: 13-57) compared to *(B) Ag Only*. For both incremental outcomes, the order of algorithm favorability remained constant across prevalence levels; however, the absolute differences between algorithms shrank as prevalence increased.

## Discussion

In this analysis, we utilized a decision analysis approach to provide a quantitative comparison of different strategies for the use of antigen tests and NAATs in SARS-CoV-2 testing programs. The six algorithms evaluated reflect differing priorities for NAAT versus antigen testing usage in populations based on resources, SARS-CoV-2 prevalence, and tolerance for missed cases and false positives. Multiple reports have found that antigen tests are less sensitive than NAATs^12-18^ and will consequently result in some antigen false-negative results among cases. The *(A) NAAT Only* and *(D) NAAT Confirmation for Ag-*neg algorithms maximize the use of NAATs to confirm negative antigen results and yielded the smallest numbers of missed cases. However, use of confirmatory NAATs for negative antigen results also incurred a need for high NAAT capacity. A strategy that selectively confirms negative antigen results with NAAT was found to be the most efficient use of a limited number NAATs [*(C) NAAT Confirmation for Sx/Ag-neg & Asx/Ag-pos*]. When uninfected people are erroneously diagnosed with SARS-CoV-2 infection (due to false-positive results), consequent isolation orders and case investigations result in lost productivity and unnecessary use of limited public health resources, and, when resulting in co-isolation with true cases, puts them at risk for ongoing exposure. Therefore, algorithms which maximize the use of NAATs to confirm positive antigen results yielded the smallest numbers of false-positive diagnoses [*(A) NAAT Only* and *(F) NAAT for Asx & Sx/Ag-pos*]. NAATs are often more costly to perform than antigen tests and may require extensive logistic arrangements for timely off-site transport and testing; also, quarantining while waiting for NAAT results can result in lost productivity. Strategies which minimize the use of NAATs [*(B) Ag Only* and *(E) Repeat Ag for Ag-neg*] offer benefits for resource-limited testing programs or in settings where lost productivity must be minimized. Each of these algorithms may be advisable depending on the programmatic goals and resource limitations of community-based SARS-CoV-2 testing programs.

Our analysis provides a quantitative framework for public health practitioners, policymakers, and stakeholders who are planning, implementing, or evaluating community-based testing programs. A reference guide discussing and applying the results of our analyses to programmatic decisions, along with key priorities, benchmarks, and indicators, is included in Supplementary Table 2. For programs intended to minimize missed cases, algorithms *(A) NAAT Only, (C) NAAT Confirmation for Sx/Ag-neg & Asx/Ag-pos*, and *(D) NAAT Confirmation for Ag-neg* are most preferable; selecting between these algorithms depends on tolerance for missed cases and available NAAT capacity. For example, at 10% prevalence, *(C) NAAT Confirmation for Sx/Ag-neg & Asx/Ag-pos* is estimated to miss 1,409 cases (95% UR: 815-2,100) but save 46 NAATs (95% UR: 29-83) for each case missed. For programs intended to minimize NAAT volume, algorithms (*B) Ag Only, (C) NAAT Confirmation for Sx/Ag-neg & Asx/Ag-pos*, and *(E) Repeat Ag for Ag-neg* are most preferable; selecting between these algorithms depends on tolerance for missed cases and available NAAT and antigen test capacity. For example, at 10% prevalence, *(E) Repeat Ag for Ag-neg* is estimated to result in 550 fewer missed cases (95% UR: 301-854) but require 92,200 more antigen tests (95% UR: 91,200-93,200) than *(B) Ag Only*.

Each algorithm evaluated in this analysis is rooted in strategies currently recommended by public health organizations except for (*A) NAAT Only*, an idealized scenario serving as a quantitative baseline. Each strategy recommended is articulated with important nuances and distinctions; algorithms analyzed here are intended to be analogous to, but not exact reproductions of these strategies. Guidance from WHO and ECDC distinguishes strategies for antigen testing in communities with low prevalence and high prevalence of SARS-CoV-2 infection. In high prevalence settings, WHO recommends considering repeat antigen testing for those with negative results,^7^ analogous to *(E) Repeat Ag for Ag-neg*; ECDC indicates that negative tests should be confirmed with RT-PCR,^8^ analogous to *(D) NAAT Confirmation for Ag-*neg. In low prevalence settings following negative antigen results, WHO recommendations indicate clinical evaluation for suspect cases in lieu of confirmatory NAATs,^7^ analogous to *(B) Ag Only*; ECDC does not recommend antigen testing for asymptomatic persons and recommends confirmatory RT-PCR for symptomatic persons with positive antigen results,^8^ analogous to *(F) NAAT for Asx & Sx/Ag-pos*. CDC interim guidance recommends a unified strategy for testing across settings,^5^ analogous to *(C) NAAT Confirmation for Sx/Ag-neg & Asx/Ag-pos*.

This decision analysis approach necessarily simplifies complex factors that may impact SARS-CoV-2 testing programs, and therefore results may not be representative of all testing programs. This analysis is intended to be representative of community-based testing (where each person receives a single test) rather than facility-based serial testing (where each person is tested on a recurring basis). In serial testing approaches, people with detected infections are isolated and removed from the testing pool, decreasing the prevalence of infection within the tested population over time. As a result, if our results were applied in those settings the results would overestimate the numbers of missed cases and testing volumes in serial testing programs. This analysis also represents a population seeking testing at a steady-state of infection and does not evaluate dynamic transmission-related outcomes intrinsic to the intervention (as a consequence of detected/missed cases) or extrinsic to the setting (due to climbing/falling incidence rates in the general population). However, other reports have provided in-depth evaluations of transmission-related outcomes in serial testing programs^19^ and symptom-based testing programs.^20^ Finally, this decision analysis approach is used to estimate expected outcomes under a theoretical perfect implementation of each algorithm. Real-world practicalities present logistical challenges to algorithm implementation: for example, up to 70% of symptomatic persons with negative antigen results may decline to participate in confirmatory NAATs.^21^ As such challenges could be addressed operationally as programs improve (e.g., collecting multiple samples at the time of initial testing), we chose a parsimonious approach of perfect implementation to highlight the fundamental distinctions between testing algorithms (independent of implementation challenges).

The results of our analysis are dependent on the accuracy and generalizability of the input parameter estimates used for each simulation. Several recent reports have described the performance characteristics of several antigen tests, with comparable results across reports when stratified by testing population symptom status.^12-18^ Programs implementing antigen tests with performance characteristics substantively different from the distributions described in Table 1 are likely to have different numbers of missed cases, depending on the assay’s sensitivity. However, only one report to date has evaluated the performance of immediate repeat antigen testing;^13^ therefore the results of *(E) Repeat Ag for Ag-neg* may not be representative of settings where immediate repeat antigen testing performs with higher sensitivity. Importantly, this parameterization does not reflect the sensitivity of delayed repeat antigen testing (e.g. as recommended by ECDC for confirmation of negative results after 2-4 days when RT-PCR capacity is limited^8^). Finally, we adopted a simplifying assumption that NAATs have 100% sensitivity and specificity as NAATs are typically considered the “gold standard” for diagnosis of SARS-CoV-2 infection. However, not all NAATs may perform similarly for confirmatory testing, and NAATs may have lower sensitivity early in the course of infection^22^ and remain positive during a patient’s post-infectious recovery.^23^ Therefore, in our approach the prevalence among persons seeking testing is representative of currently and recently infected persons detectable by high-sensitivity NAATs at the time of testing (comparable to field studies which report performance characteristics of antigen tests). Similarly, some “missed cases” in this approach are likely to represent post-infectious persons who are no longer detectable by antigen tests but remain detectable by NAAT.

Our results provide the first quantitative comparison of the expected performance of different strategies for community-based SARS-CoV-2 testing programs recommended by public health organizations. None of the algorithms evaluated in this analysis is likely to be optimal in all settings and for all programmatic priorities, and this analysis provides a framework for selecting setting-specific strategies to achieve an acceptable balance and trade-offs between programmatic priorities and constraints. As global responses to the COVID-19 pandemic continue to evolve and adapt, our results contribute to the body of evidence that will help inform SARS-CoV-2 testing strategies.

## Data Availability

Code for the algorithm simulations can be found on the CDC Epidemic Prediction Initiative GitHub site (https://github.com/cdcepi).

## Acknowledgements

The authors would like to thank the contributions of members of the Centers for Disease Control and Prevention’s Coronavirus Disease 2019 (COVID-19) Response Team: John Paul Bigouette, Lauren Boyle-Estheimer, Dustin Currie, Juliana DaSilva, Alicia Fry, Aron Hall, Michael Iademarco, Michael Johansson, Emiko Kamitani, Sandor Karpathy, Marie Killerby, Nancy Knight, Shirley Lecher, Kaitlin Mitchell, Clint Morgan, Michelle O’Hegarty, Prabasaj Paul, Reynolds Salerno, Hannah Segaloff, Rachel Slayton, Tarah Somers, Miriam Van Dyke, and Melissa Whaley.

## Footnotes

§ *See e*.*g*., *45 C.F.R. part 46.102(l)(2), 21 C.F.R. part 56; 42 U.S.C. §241(d); 5 U.S.C. §552a; 44 U.S.C. §3501 et seq*.

