## Supplementary Materials for "Quantitative Comparison of SARS-CoV-2 Nucleic Acid Amplification Test and Antigen Testing Algorithms: A Decision Analysis Simulation Model"

### Supplementary Results

*Sensitivity Analyses*

Parameters strongly correlated with the number of missed cases under each algorithm are depicted using PRCCs in Supplementary Figure 1 (assumptions of monotonicity were validated in Supplementary Figure 2). Four key parameters drove variation in the number of missed cases across algorithms. The prevalence of symptoms among infected persons was the only parameter strongly correlated with missed cases in every algorithm; it was positively correlated with missed cases in the *(F) NAAT Confirmation for Asx & Sx/Ag-pos* algorithm (where it determines how many cases receive antigen testing instead of NAAT) and was inversely correlated with missed cases in all other algorithms (where initial antigen testing is more sensitive among symptomatic persons). In each algorithm, strong inverse correlations were observed for number of missed cases and antigen test sensitivity among symptomatic persons, asymptomatic persons, or both (depending on whether these populations receive NAAT after antigen testing). Given the observed strong correlations for antigen sensitivities in symptomatic and asymptomatic persons, two-way sensitivity analyses were performed to illustrate the impact of independent variations in these parameters and the prevalence of symptoms among cases (Supplementary Figures 3-6).

### Supplementary Figures and Table

**Supplementary Figure 1. Partial Rank Correlation Coefficients (PRCCs) of Parameters Correlated with Missed Cases**

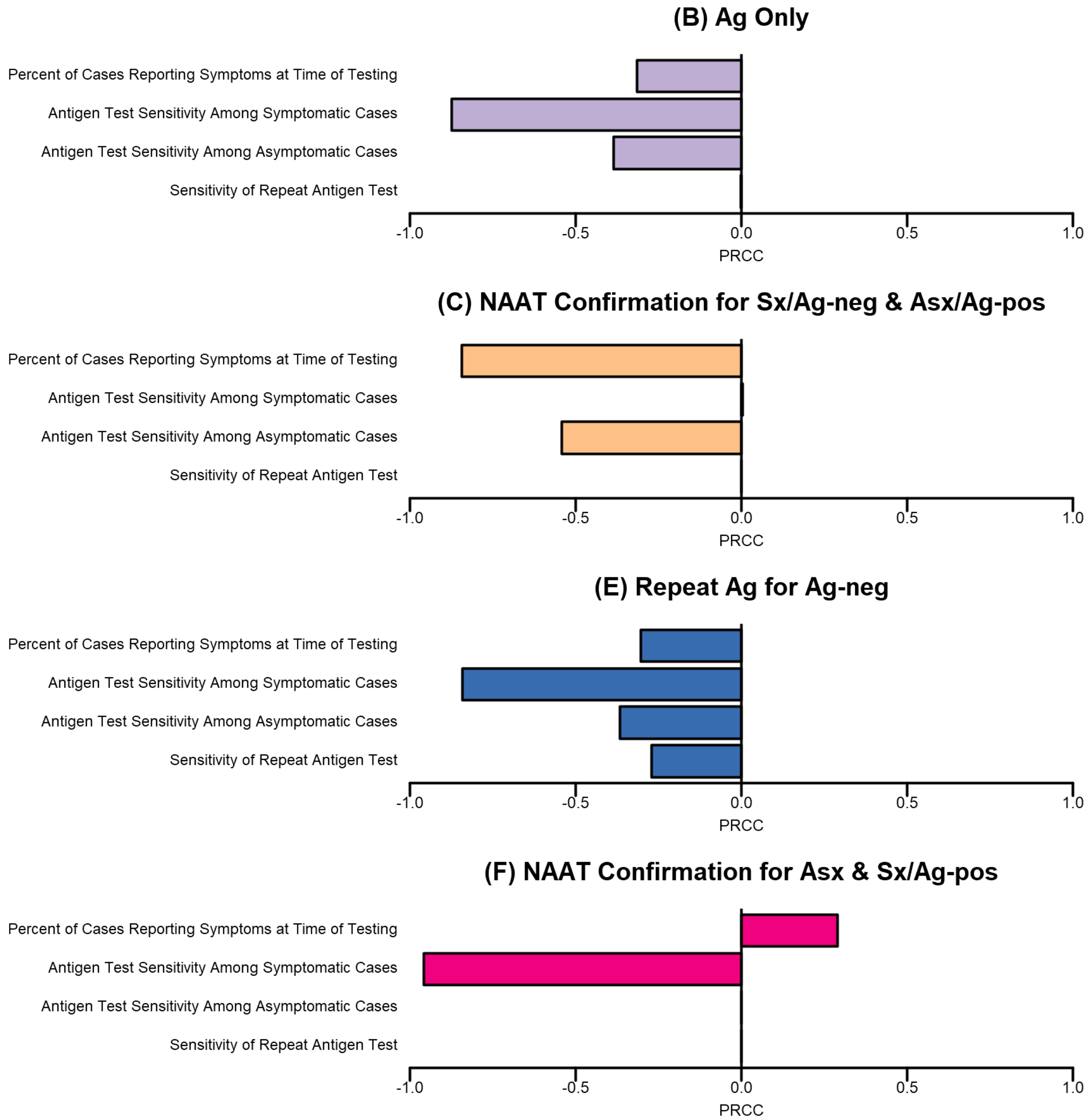


Parameters positively and negatively correlated with missed cases across 50,000 simulations are illustrated for four algorithms. PRCCs close to 1.0 reflect strong positive correlations between the parameter's input value and the number of missed cases in the results of a simulation, after accounting for variation in all other input parameters; PRCCs close to -1.0 reflect strong negative correlations between the parameter's input value and the number of missed cases; PRCCs close to 0 reflect independence of parameter value and missed cases. Parameters very weakly correlated with missed cases (|PRCC|<0.01) in all algorithms are excluded. Algorithm abbreviations and descriptions – *(B) Ag Only*: each person tested a single antigen test; *(C) NAAT Confirmation for Sx/Ag-neg and Asx/Ag-pos***:** each person receives an antigen test and NAAT is used to confirm diagnoses in persons for whom antigen results do not match binary symptom status (e.g., a symptomatic person whose antigen result is negative); *(E) Repeat Ag Confirmation of Ag*-neg: each person receives an antigen test and, for those with initial negative results, a repeat antigen test (performed within approximately 30 minutes of the initial test) is used to confirm negative diagnoses; *(F) NAAT for Asx & Sx/Ag-pos*: – asymptomatic persons receive a NAAT, while symptomatic persons receive an antigen test followed by a NAAT for those with positive antigen results.

**Supplementary Figure 2. Assessment of Monotonicity Between Parameter Input Values and Missed Cases per 100,000.**

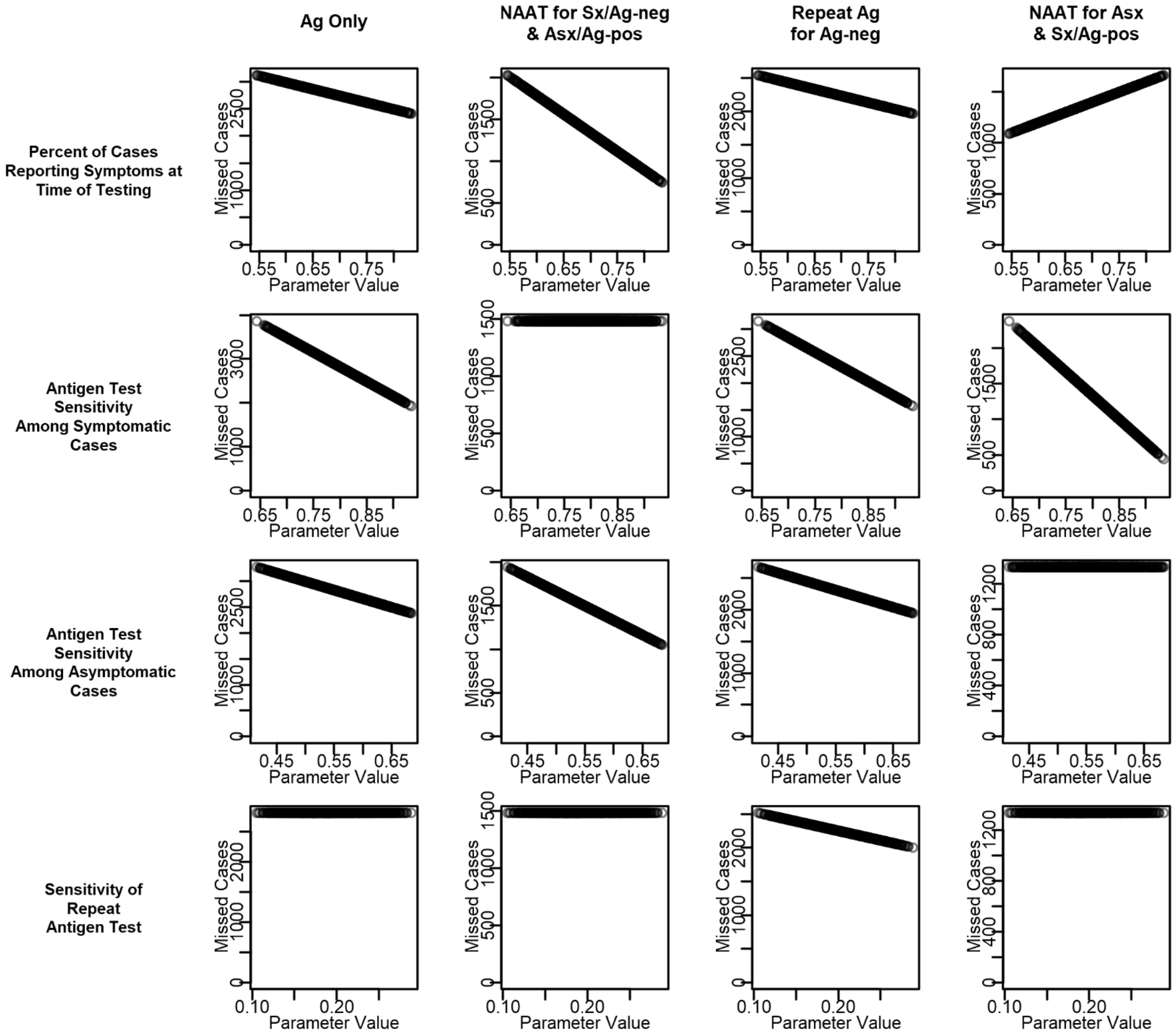


Partial Rank Correlation Coefficients (PRCCS, Supplementary Figure 1) provide valid assessments of correlations between model inputs and outcomes only if the relationship between each input parameter and the outcome is monotonic across the domain of the input parameter; this figure assesses the assumption of monotonicity. Each panel represents the relationship between one input parameter and the number of missed cases per 100,000 when simulated under one of four testing algorithms (assessed in Supplementary Figure 1). Each plot depicts the results of 1,000 simulations, with the parameter value of interest (x-axis) sampled randomly from across its domain. Within each row, all other parameters are held at their modal values in a population of 10% prevalence. Algorithm abbreviations and descriptions – *(B) Ag Only*: each person tested a single antigen test; *(C) NAAT Confirmation for Sx/Ag-neg and Asx/Ag-pos***:** each person receives an antigen test and NAAT is used to confirm diagnoses in persons for whom antigen results do not match binary symptom status (e.g., a symptomatic person whose antigen result is negative); *(E) Repeat Ag Confirmation of Ag*-neg: each person receives an antigen test and, for those with initial negative results, a repeat antigen test (performed within approximately 30 minutes of the initial test) is used to confirm negative diagnoses; *(F) NAAT for Asx & Sx/Ag-pos*: asymptomatic persons receive a NAAT, while symptomatic persons receive an antigen test followed by a NAAT for those with positive antigen results.

**Supplementary Figure 3. Missed Cases per 100,000 Persons Tested as a Function of Antigen Test Sensitivity and the Prevalence of Symptoms Among Cases in a Population of 5% SARS-CoV-2 Prevalence.**

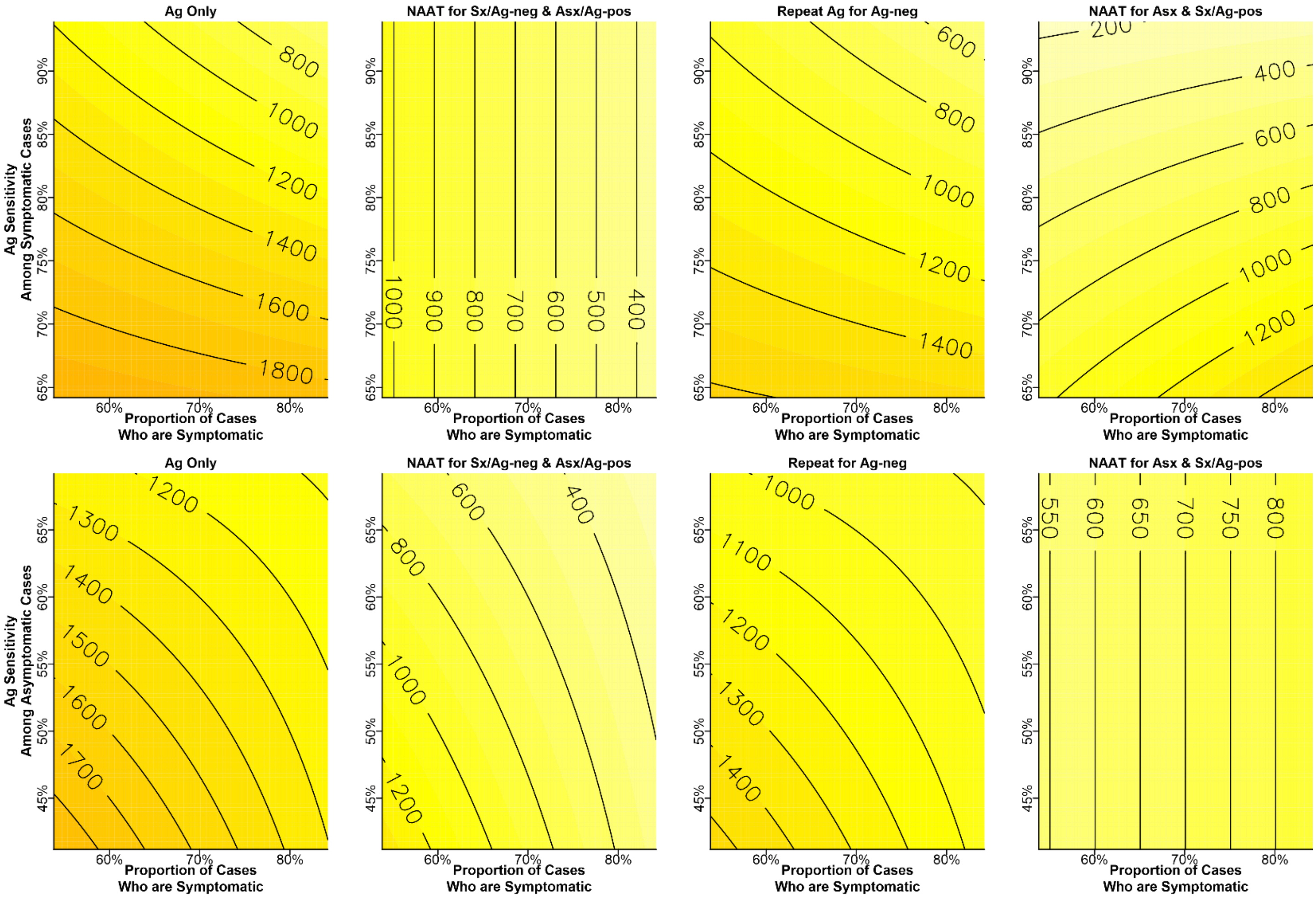


Each plot illustrates the number of cases missed at 5% prevalence in a population of 100,000 seeking testing when simulated with different combinations of input values for two parameters: the proportion of cases that are symptomatic (x-axis) and the sensitivity of antigen testing (y-axis) in either symptomatic cases (top row) or asymptomatic cases (bottom row). Colors and contour lines indicate the number of missed cases resulting from a particular combination of the two parameters. For these simulations, all other parameters were held constant at their modal values (see Table 1). Algorithm abbreviations and descriptions – *(B) Ag Only*: each person tested a single antigen test; *(C) NAAT Confirmation for Sx/Ag-neg and Asx/Ag-pos***:** each person receives an antigen test and NAAT is used to confirm diagnoses in persons for whom antigen results do not match binary symptom status (e.g., a symptomatic person whose antigen result is negative); *(E) Repeat Ag Confirmation of Ag*-neg: each person receives an antigen test and, for those with initial negative results, a repeat antigen test (performed within approximately 30 minutes of the initial test) is used to confirm negative diagnoses; *(F) NAAT for Asx & Sx/Ag-pos*: asymptomatic persons receive a NAAT, while symptomatic persons receive an antigen test followed by a NAAT for those with positive antigen results.

**Supplementary Figure 4. Missed Cases per 100,000 Persons Tested as a Function of Antigen Test Sensitivity and the Prevalence of Symptoms Among Cases in a Population of 10% SARS-CoV-2 Prevalence.**

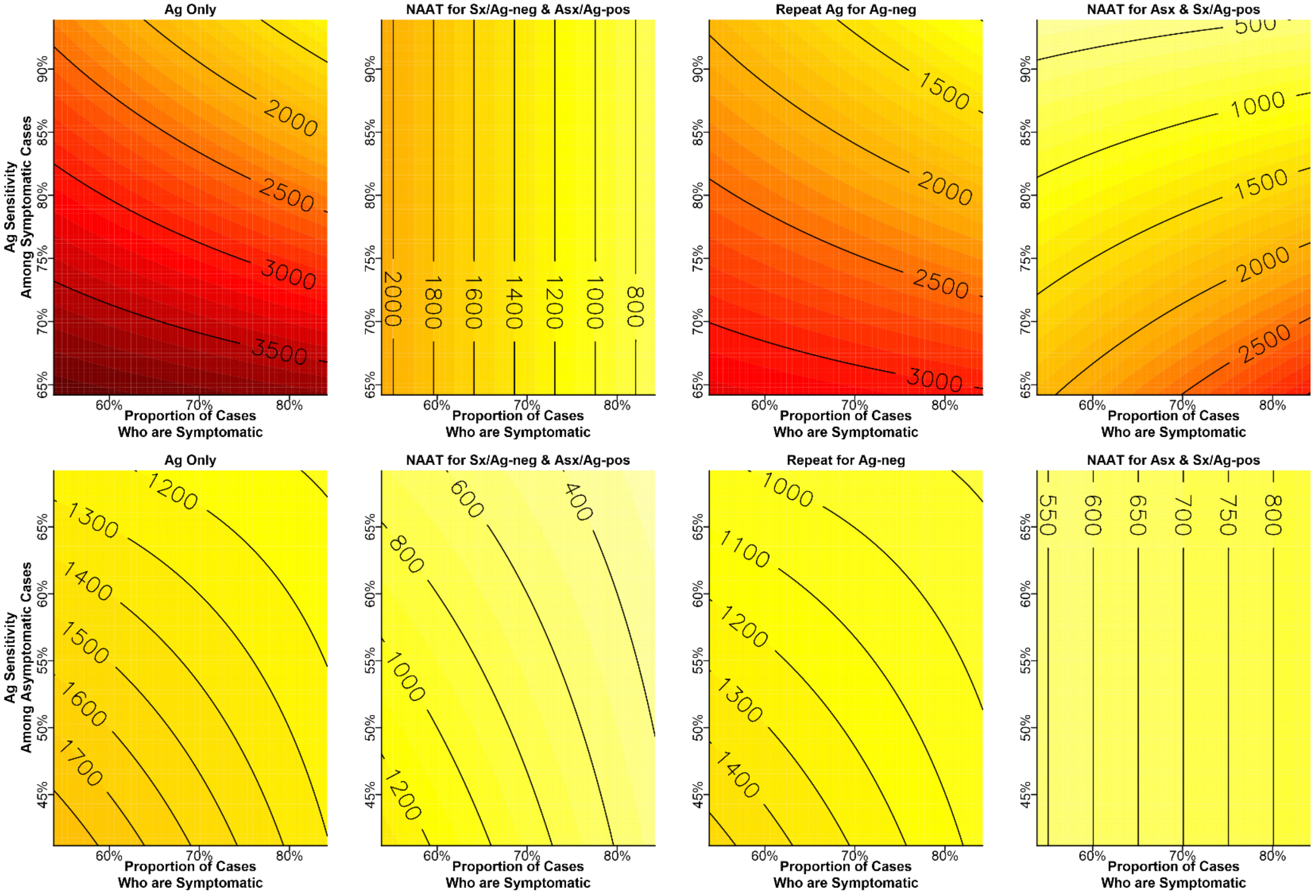


Each plot illustrates the number of cases missed at 10% prevalence in a population of 100,000 seeking testing when simulated with different combinations of input values for two parameters: the proportion of cases that are symptomatic (x-axis) and the sensitivity of antigen testing (y-axis) in either symptomatic cases (top row) or asymptomatic cases (bottom row). Colors and contour lines indicate the number of missed cases resulting from a particular combination of the two parameters. For these simulations, all other parameters were held constant at their modal values (see Table 1). Algorithm abbreviations and descriptions – *(B) Ag Only*: each person tested a single antigen test; *(C) NAAT Confirmation for Sx/Ag-neg and Asx/Ag-pos***:** each person receives an antigen test and NAAT is used to confirm diagnoses in persons for whom antigen results do not match binary symptom status (e.g., a symptomatic person whose antigen result is negative); *(E) Repeat Ag Confirmation of Ag*-neg: each person receives an antigen test and, for those with initial negative results, a repeat antigen test (performed within approximately 30 minutes of the initial test) is used to confirm negative diagnoses; *(F) NAAT for Asx & Sx/Ag-pos*: asymptomatic persons receive a NAAT, while symptomatic persons receive an antigen test followed by a NAAT for those with positive antigen results.

**Supplementary Figure 5. Missed Cases per 100,000 Persons Tested as a Function of Antigen Test Sensitivity and the Prevalence of Symptoms Among Cases in a Population of 15% SARS-CoV-2 Prevalence.**

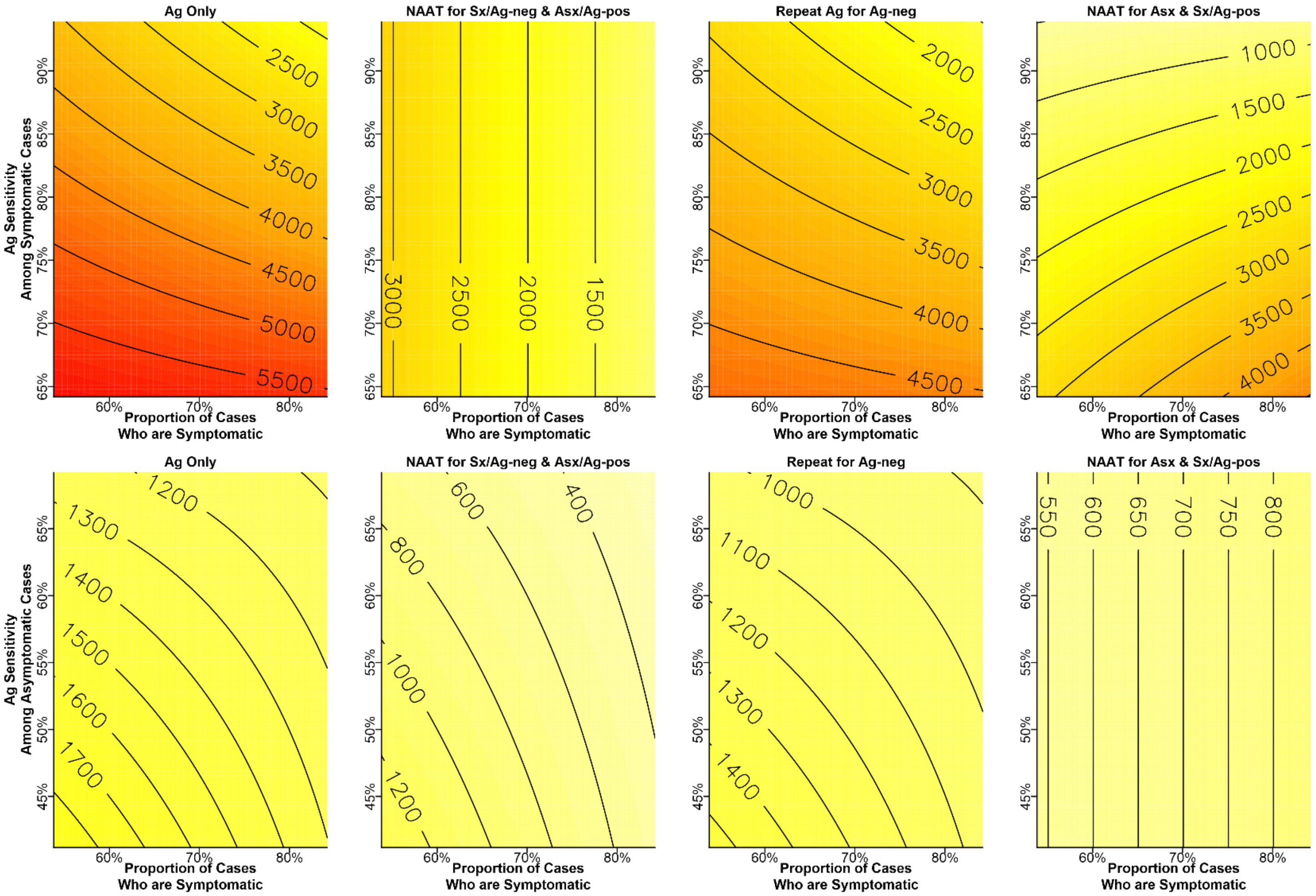


Each plot illustrates the number of cases missed at 15% prevalence in a population of 100,000 seeking testing when simulated with different combinations of input values for two parameters: the proportion of cases that are symptomatic (x-axis) and the sensitivity of antigen testing (y-axis) in either symptomatic cases (top row) or asymptomatic cases (bottom row). Colors and contour lines indicate the number of missed cases resulting from a particular combination of the two parameters. For these simulations, all other parameters were held constant at their modal values (see Table 1). Algorithm abbreviations and descriptions – *(B) Ag Only*: each person tested a single antigen test; *(C) NAAT Confirmation for Sx/Ag-neg and Asx/Ag-pos***:** each person receives an antigen test and NAAT is used to confirm diagnoses in persons for whom antigen results do not match binary symptom status (e.g., a symptomatic person whose antigen result is negative); *(E) Repeat Ag Confirmation of Ag*-neg: each person receives an antigen test and, for those with initial negative results, a repeat antigen test (performed within approximately 30 minutes of the initial test) is used to confirm negative diagnoses; *(F) NAAT for Asx & Sx/Ag-pos*: asymptomatic persons receive a NAAT, while symptomatic persons receive an antigen test followed by a NAAT for those with positive antigen results.

**Supplementary Figure 6. Missed Cases per 100,000 Persons Tested as a Function of Antigen Test Sensitivity and the Prevalence of Symptoms Among Cases in a Population of 20% SARS-CoV-2 Prevalence.**

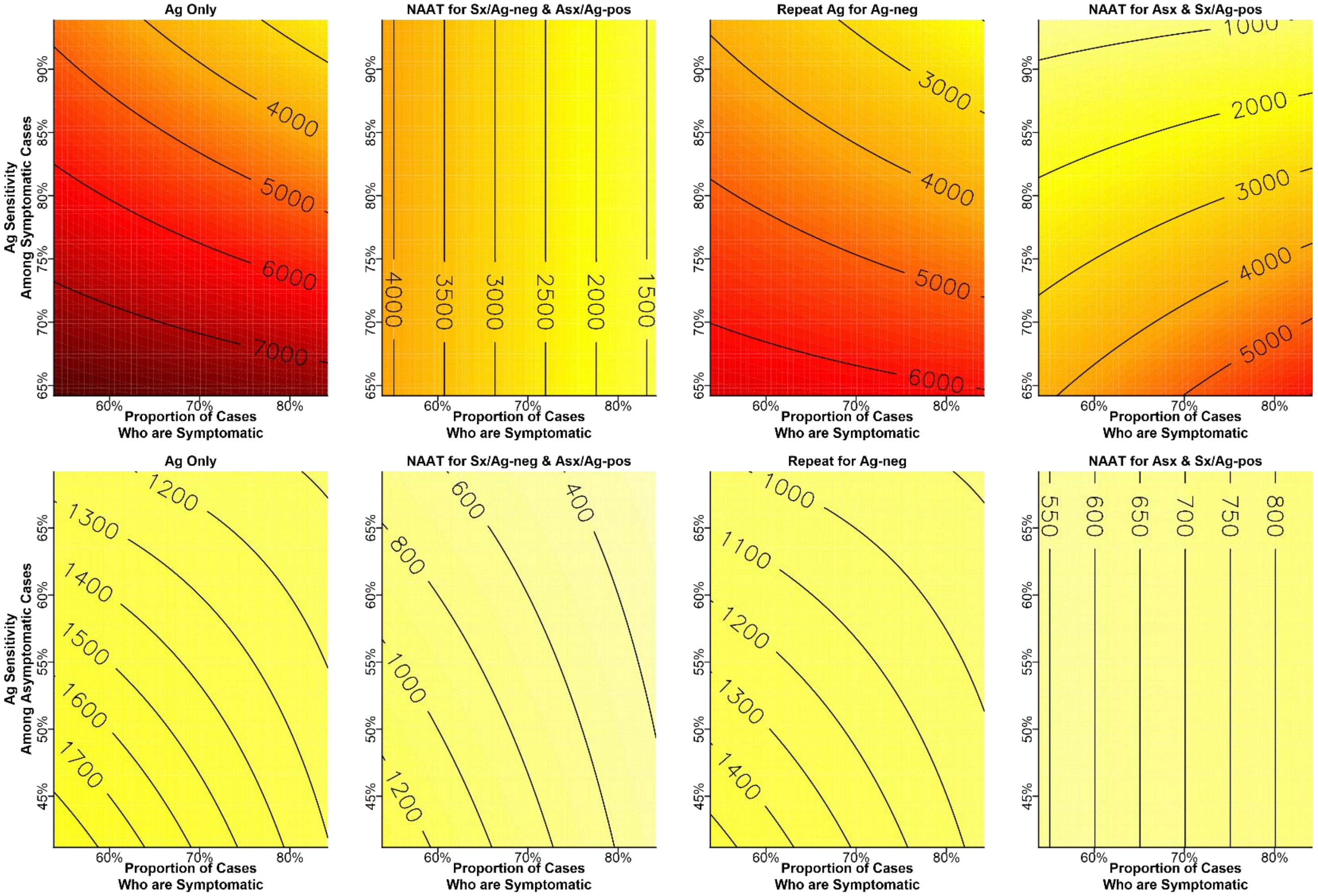


Each plot illustrates the number of cases missed at 20% prevalence in a population of 100,000 seeking testing when simulated with different combinations of input values for two parameters: the proportion of cases that are symptomatic (x-axis) and the sensitivity of antigen testing (y-axis) in either symptomatic cases (top row) or asymptomatic cases (bottom row). Colors and contour lines indicate the number of missed cases resulting from a particular combination of the two parameters. For these simulations, all other parameters were held constant at their modal values (see Table 1). Algorithm abbreviations and descriptions – *(B) Ag Only*: each person tested a single antigen test; *(C) NAAT Confirmation for Sx/Ag-neg and Asx/Ag-pos***:** each person receives an antigen test and NAAT is used to confirm diagnoses in persons for whom antigen results do not match binary symptom status (e.g., a symptomatic person whose antigen result is negative); *(E) Repeat Ag Confirmation of Ag*-neg: each person receives an antigen test and, for those with initial negative results, a repeat antigen test (performed within approximately 30 minutes of the initial test) is used to confirm negative diagnoses; *(F) NAAT for Asx & Sx/Ag-pos*: asymptomatic persons receive a NAAT, while symptomatic persons receive an antigen test followed by a NAAT for those with positive antigen results.

**Supplementary Table 1. Primary and Secondary Outcomes of SARS-CoV-2 Nucleic Acid Amplification Test (NAAT) and Antigen (Ag) Testing Algorithms per 100,000 Persons Tested**.

| Outcome* | Prevalence | *(A) NAAT Only* | *(B) Ag Only* | *(C) NAAT Confirmation for Sx/Ag-neg & Asx/Ag-pos* | *(D) NAAT Confirmation for Ag-neg* | *(E) Repeat Ag for Ag-neg* | *(F) NAAT Confirmation for Asx & Sx/Ag-pos* |
|---|---|---|---|---|---|---|---|
| **Missed Cases*** | **5%** | 0 | 1415 (945-1870) | 705 (407-1050) | 0 | 1140 (753-1534) | 694 (311-1140) |
| | **10%** | 0 | 2830 (1890-3740) | 1409 (815-2100) | 0 | 2280 (1507-3067) | 1389 (622-2280) |
| | **15%** | 0 | 4244 (2835-5611) | 2114 (1222-3150) | 0 | 3420 (2260-4601) | 2083 (933-3420) |
| | **20%** | 0 | 5659 (3780-7481) | 2819 (1629-4200) | 0 | 4561 (3014-6135) | 2777 (1244-4560) |
| **False Positive Diagnoses*** | **5%** | 0 | 671 (328-1089) | 141 (29-348) | 671 (328-1089) | 738 (381-1166) | 0 |
| | **10%** | 0 | 635 (311-1031) | 134 (27-330) | 635 (311-1031) | 699 (361-1105) | 0 |
| | **15%** | 0 | 600 (294-974) | 127 (26-311) | 600 (294-974) | 660 (341-1043) | 0 |
| | **20%** | 0 | 565 (276-917) | 119 (24-293) | 565 (276-917) | 621 (321-982) | 0 |
| **NAAT Volume*** | **5%** | 100000 | 0 | 34073 (22486-48110) | 95735 (95113-96320) | 0 | 67338 (53175-79019) |
| | **10%** | 100000 | 0 | 33957 (22963-47262) | 92183 (91165-93163) | 0 | 68324 (54892-79399) |
| | **15%** | 100000 | 0 | 33819 (23381-46441) | 88632 (87177-90043) | 0 | 69296 (56574-79795) |
| | **20%** | 100000 | 0 | 33712 (23749-45646) | 85083 (83175-86935) | 0 | 70257 (58257-80257) |
| **Antigen Test Volume*** | **5%** | 0 | 100000 | 100000 | 100000 | 195735 (195113-196320) | 35504 (23764-49741) |
| | **10%** | 0 | 100000 | 100000 | 100000 | 192183 (191165-193163) | 37222 (26071-50761) |
| | **15%** | 0 | 100000 | 100000 | 100000 | 188632 (187177-190043) | 38966 (28329-51808) |
| | **20%** | 0 | 100000 | 100000 | 100000 | 185083 (183175-186935) | 40737 (30504-52866) |
| **Person-Days of Unnecessary Quarantine Awaiting NAAT** | **5%** | 145943 (67713-242315) | 0 | 95817 (42477-174390) | 144996 (67289-240922) | 0 | 48103 (18455-98293) |
| | **10%** | 138262 (64149-229561) | 0 | 90774 (40242-165211) | 137365 (63748-228242) | 0 | 45571 (17484-93119) |
| | **15%** | 130581 (60585-216808) | 0 | 85731 (38006-156033) | 129733 (60206-215562) | 0 | 43040 (16512-87946) |
| | **20%** | 122900 (57021-204055) | 0 | 80688 (35771-146854) | 122102 (56665-202882) | 0 | 40508 (15541-82773) |
| **Ratio of Saved NAATs to Additional Missed Cases†** | **5%** | NA | 71 (53-106) | 93 (58-165) | Positive Infinity§ | 88 (65-133) | 47 (24-112) |
| | **10%** | NA | 35 (27-53) | 46 (29-83) | Positive Infinity§ | 44 (33-66) | 23 (12-53) |
| | **15%** | NA | 24 (18-35) | 31 (20-55) | Positive Infinity§ | 29 (22-44) | 15 (8-34) |
| | **20%** | NA | 18 (13-26) | 23 (15-42) | Positive Infinity§ | 22 (16-33) | 11 (6-24) |
| **Ratio of Added NAATs to Additional Detected Cases ‡** | **5%** | 71 (53-106) | NA | 49 (26-116) | 68 (51-101) | 0 | 95 (60-168) |
| | **10%** | 35 (27-53) | NA | 25 (13-57) | 33 (25-48) | 0 | 48 (31-85) |
| | **15%** | 24 (18-35) | NA | 16 (9-37) | 21 (16-31) | 0 | 33 (21-57) |
| | **20%** | 18 (13-26) | NA | 12 (7-27) | 15 (12-22) | 0 | 25 (16-43) |

* All results are presented as median (95% uncertainty range). Results presented with no uncertainty range indicate that this outcome is not a function of the sampled parameter values and is fixed at either 0 or the population size (100,000) for all simulations of this algorithm.
† Compared to the *(A) NAAT Only* algorithm; the results of this outcome for this algorithm is listed as NA (not applicable).
‡ Compared to the *(B) Ag Only* algorithm; the results of this outcome for this algorithm is listed as NA (not applicable).
§ Missed cases under this algorithm are 0 and saved NAATs are greater than zero; therefore, this ratio is always mathematically equal to positive infinity.

**Supplementary Table 2. Decision Analysis Results Reference Guide**

<table>
<tr><th>Priority Metric</th><th>Algorithms* to Consider</th><th>Pros†</th><th>Cons†</th><th>Synthesis</th><th>Impact of Prevalence</th></tr>
<tr><td rowspan="3">Minimal Missed Cases</td><td>(A) NAAT Only</td><td>1. No missed cases<br>2. No false positives<br>3. No need for Ag testing infrastructure</td><td>1. Highest NAAT volume<br>2. Highest unneeded quarantine while waiting for results</td><td rowspan="3">Testing all persons with NAAT or confirming all Ag- results with NAAT ensures no cases are missed.<br><br>(D) requires between 96% NAAT volume (at 5% prevalence) and 85% NAAT volume (at 20% prevalence) plus 100% Ag test capacity.<br><br>(F) misses 14% of cases relative to (A) and (D), but saves between 47 NAATs per missed case (at 5% prevalence) and 11 NAATs per missed case (at 20% prevalence).</td><td rowspan="3">At low prevalence, cases are rare and many NAATs are needed for each case detected in (A) and (D).<br><br>As prevalence increases, the absolute number of missed cases from (F) increases.<br><br>Minimal missed cases from (A) and (D) becomes more favorable and (F) becomes less favorable.</td></tr>
<tr><td>(D) NAAT for Ag-neg</td><td>1. No missed cases</td><td>1. High false-positives<br>2. High NAAT volume<br>3. High Ag volume<br>4. High unneeded quarantine while waiting for results</td></tr>
<tr><td>(F) NAAT for Asx & Sx/Ag-pos</td><td>1. Low missed cases<br>2. No false positives<br>3. Low unneeded quarantine while waiting for results<br>4. Low Ag volume</td><td>1. Moderate NAAT volume</td></tr>
<tr><td rowspan="3">Minimal NAAT Capacity</td><td>(B) Ag Only</td><td>1. No NAAT infrastructure required<br>2. No unneeded quarantine while waiting for results</td><td>1. Highest missed cases<br>2. High false-positives<br>3. High Ag volume</td><td rowspan="3">Eliminating NAAT/confirmation provides faster results, at the expense of more incorrect results (missed cases and false positives).<br><br>(E) reduces missed cases by 19% relative to (B), but requires between 96% (at 5%) and 85% (at 20% prevalence) greater Ag test volume relative to (B) and (C).<br><br>(C) requires 34% NAAT test volume and will require between 49 NAATs (at 5% prevalence) and 12 NAATs (at 20% prevalence) for each additional case detected, relative to (B).</td><td rowspan="3">At low prevalence, cases are fewer and more Ag- results need NAAT under (C).<br><br>As prevalence increases, NAAT volume decreases with (C); it remains 0 for (B) and (E). Simultaneously, absolute missed cases increase more under (B) and (E).<br><br>As prevalence increases, negative consequences of (B) and (E) become less favorable and (C) becomes more favorable.</td></tr>
<tr><td>(E) Repeat Ag for Ag-neg</td><td>1. No NAAT infrastructure required<br>2. No unneeded quarantine while waiting for results</td><td>1. High missed cases<br>2. Highest false-positives<br>3. Highest Ag volume</td></tr>
<tr><td>(C) NAAT for Sx/Ag-neg & Asx/Ag-pos</td><td>1. Low NAAT volume<br>2. Moderate missed cases<br>3. Low false positives</td><td>1. Moderate unneeded quarantine while waiting for results<br>2. High Ag volume</td></tr>
</table>

*See Methods and Figure 1 for full descriptions of each algorithm evaluated.

† Except where stated otherwise, numerical results are simplified by rank order for summary as follows: *Highest* refers to the algorithm for which the outcome is the highest number (compared to all other algorithms, across prevalence levels); *High* refers to algorithms which result in the second- or third-highest level outcome of algorithms evaluated; *Moderate* refers to the middle level of outcome (when outcomes from multiple algorithms are equal); *Low* refers to algorithms with result in the second- or third-lowest level of outcome; *Lowest* refers to the algorithm for which the outcome is lowest (when this lowest level is zero, this is stated as, e.g., "No missed cases"). See Results and Figure 2 for exact numerical results.

Abbreviations: NAAT – nucleic acid amplification tests (such as RT-PCR); Ag – antigen; Ag-pos - positive antigen result; Ag-neg - negative antigen result.

**Supplementary Table 2 (Continued)**

| Priority Metric | Algorithms* to Consider | Pros† | Cons† | Synthesis | Impact of Prevalence |
|---|---|---|---|---|---|
| Minimal False-positives | *(A) NAAT Only* | 1. No false positives<br>2. No missed cases<br>3. No Ag testing infrastructure required | 1. Highest NAAT volume<br>2. Highest level of unneeded quarantine while waiting for results | Testing all persons with NAAT or confirming all Ag+ results with NAAT ensures no false positives are issued.<br><br>(F) requires between 67% NAAT volume (at 5% prevalence) and 70% NAAT volume (at 20% prevalence) and 36% to 41% Ag testing volume (respectively), but results in between 14% of cases missed relative to A).<br><br>(C) requires 34% NAAT volume and results in 14% of cases missed relative to (A). | At low prevalence, cases are rare and false-positives are greatest for C); they are 0 for (A) and (F).<br><br>As prevalence increases, absolute false-positives decrease for (C).<br><br>As prevalence increases, the benefits of (C) become more favorable. |
| | *(F) NAAT for Asx & Sx/Ag-pos* | 1. No false positives<br>2. Low missed cases<br>3. Low unneeded quarantine while waiting for results<br>4. Low Ag volume | 1. Moderate NAAT volume | | |
| | *(C) NAAT for Sx/Ag-neg & Asx/Ag-pos* | 1. Low false positives<br>2. Moderate missed cases<br>3. Low NAAT volume | 1. Moderate unneeded quarantine while waiting for results<br>2. High Ag volume | | |
| Balance between Missed Cases and NAAT Volume | *(C) NAAT for Sx/Ag-neg & Asx/Ag-pos* | 1. Moderate missed cases<br>2. Low NAAT volume<br>3. Low false positives | 1. Moderate unneeded quarantine while waiting for results<br>2. High Ag volume | (D) results in no missed cases and saves between 4% NAAT test volume (at 5% prevalence) and 15% NAAT test volume (at 20% prevalence) relative to (A).<br><br>(C) results in more missed cases but greatly reduces NAAT test volume (by 66%) compared to (A). This will save between 93 (at 5% prevalence) and 93 (at 20% prevalence) NAATs for each additional case missed.<br><br>(E) eliminates NAAT entirely but substantially increase missed cases (23% compared to (A)). This will save between 87 (at 5% prevalence) and 22 (at 20% prevalence) NAATs for each additional case missed. | At low prevalence, cases are rare and many NAATs are needed for each case detected in (C), (D) and (E).<br><br>As prevalence increases, cases increase more than NAAT volume increases and fewer NAATs are needed for each case detected in (C), (D) and (E). Absolute numbers of missed cases increase more and (E) than (C) (and remain 0 for (A) and (D)).<br><br>As prevalence increases, the efficiency of (C), (D), and (E) becomes more favorable, while the negative consequences of (C) and (E) become less favorable. |
| | *(D) NAAT for Ag-neg* | 1. No missed cases | 1. High false-positives<br>2. High NAAT volume<br>3. High Ag volume<br>4. High unneeded quarantine while waiting for results | | |
| | *(E) Repeat Ag for Ag-neg* | 1. No NAAT infrastructure required<br>2. No unneeded quarantine while waiting for results | 1. High missed cases<br>2. Highest false-positives<br>3. Highest Ag volume | | |
| | *(A) NAAT Only* | 1. No missed cases<br>2. No false positives<br>3. No need for Ag testing infrastructure | 1. Highest NAAT volume<br>2. Highest unneeded quarantine while waiting for results | | |
